# Reduction of perioperative rebleeding in aneurysmal subarachnoid hemorrhage with combination therapy of hemocoagulase and antifibrinolytics: A multicenter retrospective study

**DOI:** 10.1101/2025.05.30.25328622

**Authors:** Sijie Sun, Jun Ma, Jiayi Li, Zhen Liu, Sitong Cui, Qingquan Li, Hongwei Fan, Runze Qiu, Yingbin Li

## Abstract

**Background:** Rebleeding following aneurysmal subarachnoid hemorrhage (aSAH) interventions significantly increases mortality and disability. While hemostatic agents are commonly used to prevent rebleeding, the efficacy of antifibrinolytics as monotherapy remains controversial, and thrombins has limited supporting evidence. This study investigates the impact of hemocoagulase, a thrombin-like agent, and its combination with antifibrinolytics on reducing rebleeding and improving functional outcomes in aSAH patients undergoing invasive treatments, providing insights into optimal medication strategies for aSAH management.

**Methods:** We conducted a retrospective analysis of data from aSAH patients across three medical institutions over a ten-year period. We compared post-treatment outcomes and the incidence of adverse events between two cohorts those who received hemocoagulase and antifibrinolytics during the perioperative period and those who did not. Primary outcomes included mortality, dependency in activities of daily living, and length of stay. Secondary outcomes comprised rebleeding and cerebral ischemic incidents.

**Results:** The study included 345 aSAH patients who underwent neurosurgical interventions, with 81.16% receiving hemostatic agents. Patients treated with hemostatic medications exhibited a significantly lower incidence of hemiplegia compared to the control cohort. However, no differences were observed in rebleeding or cerebral ischemic events. Notably, for patients who received a combination of hemocoagulase and a single antifibrinolytic agent, the incidence of rebleeding within 72 hours post-intervention and overall postoperative rebleeding was significantly lower.

**Conclusion:** The use of hemostatic agents during the perioperative period of aSAH may improve functional outcomes. Combining hemocoagulase with a single antifibrinolytic agent appears to reduces early rebleeding following aSAH interventions.

## Introduction

Aneurysmal subarachnoid hemorrhage (aSAH) accounts for 5% of all stroke cases and is associated with a mortality rate of 32% to 67%. One-third of patients face poor outcomes[1,2]. aSAH can lead to early brain injury (EBI) characterized by vasospasm, microthrombus formation, and disruption of the blood-brain barrier. These pathophysiological changes ultimately result in delayed cerebral ischemia (DCI)[3], which typically occurs 4 to 10 days post-aSAH as an independent predictor of poor functional and cognitive outcomes[3,4].

Early rebleeding occurring within 72 hours in 8-23% of aSAH cases, with 9-17% happening within the first 24 hours[5], significantly increases the risk of poor outcomes. Mortality rates for patients who experience early rebleeding can be up to 50%-80%[6,7]. Rebleeding is often associated with untreated aneurysms or delayed treatment[8], underscoring the importance of timely neurosurgical interventions to prevent recurrent bleeding[9]. However, not all aSAH patients can receive interventions immediately upon admission, and in such cases, short-term use of hemostatic agents may delay rebleeding, providing a critical window for risk control[1,10–12].

As traditional hemostatic agents, antifibrinolytics exert a blood-clotting effect but also increase the risk of ischemic complications. Studies have shown that perioperative use of antifibrinolytics alone does not reduce the risk of rebleeding[13–15] and has not improved overall outcomes in aSAH patients[11,15–17]. Hemocoagulase, derived from detoxified and purified snake venom, is a thrombin-like enzyme that accelerates fibrin clot formation without inducing factor XIII production [18]. It is characterized by low toxicity, rapid onset, prolonged action, and a lack of intravascular thrombosis induction. Importantly, its hemostatic effects are not antagonized by heparin[19,20], making it a promising agent for preventing surgical bleeding [19,21,22]. Despite the widespread use of hemocoagulase in various clinical settings, preliminary research suggests only a potential neuroprotective effect in experimental models of traumatic brain injury[23]. Robust clinical evidence for preventing rebleeding in aSAH is still lacking.

Studies have demonstrated a synergistic hemostatic effect between thrombin-like compounds and tranexamic acid, significantly reducing perioperative bleeding in femoral intertrochanteric fracture repair[24]. However, this combination did not provide the anticipated hemostatic benefits in orthognathic surgery[25], highlighting the need for further investigation into its efficacy in different clinical contexts. Given these findings, the question of whether the combination of hemocoagulase and antifibrinolytics can reduce early rebleeding and improve outcomes of aSAH patients following invasive treatments remains an area of active investigation.

To address the current gaps, we conducted a 10-year multicenter retrospective cohort study to evaluate the impact of hemocoagulase, both as monotherapy and in combination with antifibrinolytics, on clinical outcomes and the incidence of perioperative rebleeding and other adverse events in aSAH patients receiving neurosurgical interventions. Additionally, this study aimed to explore how different administration strategies influence the incidence of adverse events and patient outcomes.

## Methods

### Patients and Settings

This retrospective cohort study was conducted across three medical institutions, Nanjing First Hospital, Nanjing Medical University, The Second Affiliated Hospital of Nanjing Medical University, and Sir Run Run Hospital, Nanjing Medical University. The study included adult patients (age ≥ 18 years) admitted between January 1 2012 and December 31 2022 who were diagnosed with SAH or ruptured intracranial aneurysm (RIA) via computed tomography angiography (CTA), digital subtraction angiography (DSA), or magnetic resonance angiography (MRA). Patients were excluded if they did not receive any form of interventional embolization or microsurgery clipping during hospitalization, were diagnosed with traumatic SAH, moyamoya disease, arteriovenous malformation, or pseudoaneurysm, chose to be discharged during treatment, or had severely incomplete clinical information.

The study was carried out in strict adherence to the Declaration of Helsinki and the requirements of the participating institutions. Ethical approval for the study protocol was obtained from the ethics committees of all participating institutions (grant numbers KY20230424-01-KS-01, 2023-KY-147-01, and 2023-SR-020). Informed consent was waived due to the retrospective nature of the study.

The analyzed characteristics included age, sex, smoking, alcohol consumption, comorbidities, Glasgow Coma Scale (GCS) on admission, aneurysm characteristics, treatment modalities, and medication strategies. Aneurysms characteristics included size, location, and multiplicity. Treatment modalities comprised microsurgical clipping and embolization coiling. Medication strategies encompassed the variety, timing, and combinations of hemostatic agent administration. In this research, the perioperative hemostatic medication regimens for patients were determined by physicians and classified into four categories: the control group, the hemostatic medications group, the single-agent group, the dual-agent group, and the triple-agent group. The single-agent group was further subdivided into the hemocoagulase subgroup (Hemocoagulase-only group) and the antifibrinolytic subgroup (Antifibrinolytic-only group). Similarly, the dual-agent group was categorized into the hemocoagulase combined with antifibrinolytic subgroup (Hemocoagulase & single antifibrinolytic group) and the dual antifibrinolytics subgroup (Dual antifibrinolytics group).

### Outcome Assessment

The primary outcomes were assessed with length of stay (LOS), adverse endpoints, and mortality. Adverse endpoints were defined as death or dependence on activities of daily living, as manifested by Glasgow Outcome Scale (GOS) 1-3, modified Rankin Scale (mRS) 4-6, or a clinical diagnosis of hemiplegia. Perioperative rebleeding and ischemia were recorded as secondary outcomes. Additionally, we collected other significant complications, such as increased intracranial pressure (IICP), hydrocephalus, cerebral vasospasm (CVS), intracranial infection, seizures, brain herniation, brain edema, fluid and electrolyte disturbances, pulmonary infection, hyperglycemia, cardiac insufficiency, hypoproteinemia, optic nerve injury, fever, organ failure, and shock.

### Statistical analysis

Categorical variables were summarized as counts and percentages and analyzed by Chi-square or Fisher’s exact test, as appropriate. Continuous variables were first tested for normality. Those following a normal distribution were expressed as mean and standard deviation (SD) and compared using Student’s t-test. For variables with non-normal distributions, medians and interquartile ranges (IQR, 25% to 75%) were provided, and comparisons were conducted using the Mann-Whitney U test. To account for baseline differences, multivariate logistic regression was employed. A two-tailed p-value < 0.05 or a 95% confidence interval (CI) not encompassing 1.0 was considered statistically significant. All statistical analysis were performed using GraphPad prism 9.0.0 (GraphPad Software, USA).

## Results

### Population Characteristics

Among 1,164 adult patients diagnosed with SAH or RIA, a total of 345 met the inclusion criteria for our study. Of these, 280 (81.16%) received hemostatic therapy, while 65 (18.84%) did not. The only difference in baseline characteristics between the two cohorts was in the intervention approach (odds ratio [OR] 0.14, 95% CI 0.05-0.32). Among the 280 patients who received hemostatic agents, 141 (50.36%) were treated with a single drug, 118 (42.14%) received a combination of two drugs, and 21 (7.50%) were prescribed a combination of three drugs. Drug combinations could be administered either simultaneously or at different times. Among the patients prescribed a single hemostatic agent, 57 (40.43%) were treated exclusively with hemocoagulase, while 84 (59.57%) received only antifibrinolytics. For those on a combination of two agents, 113 (95.76%) were given hemocoagulase in conjunction with an antifibrinolytic agent, and the remaining 5 (4.24%) received a combination of two different antifibrinolytic agents. All cases where three hemostatic agents were used concurrently involved hemocoagulase along with two antifibrinolytic agents **(Figure 1)**.

**Figure 1.**
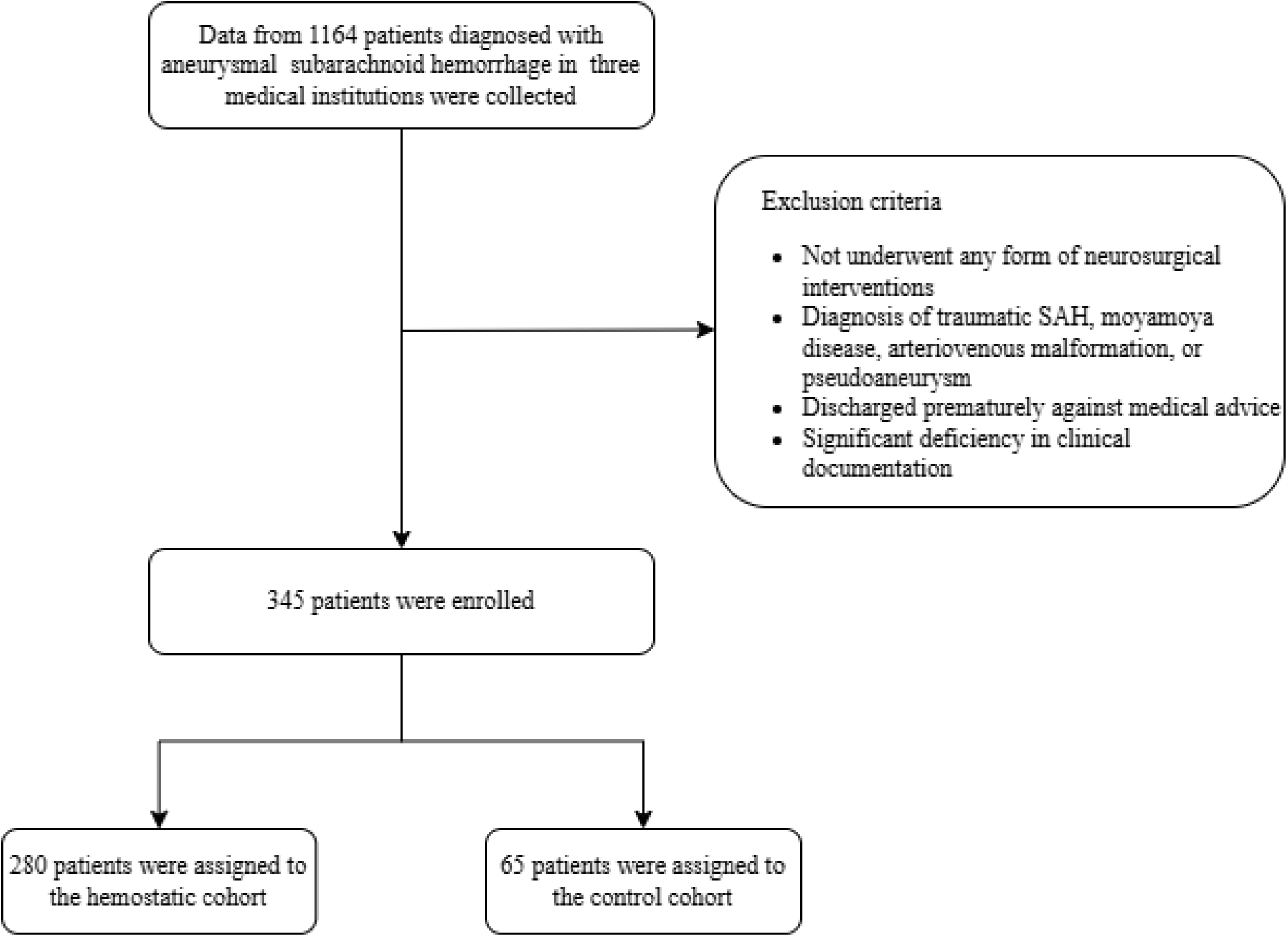
Screening and enrolling profile.

Among patients receiving various hemostatic medication regimens, the proportion treated with interventional embolization or clipping differed significantly compared to those not using hemostatic agents (Hemostatic medications: OR 0.14, 95% CI 0.05-0.32; Single-agent: OR 3.67, 95% CI 1.47-11.15; Dual-agent: OR 9.78, 95% CI 3.99-29.54; Triple-agent: OR 16.00, 95% CI 4.81-61.37). Patients in the single-agent cohort were slightly younger than those not received hemostatic medications (60.67 ± 10.13 years vs 61.54 ± 12.29 years, OR 0.87, 95% CI 0.52-0.38). The dual-agent cohort exhibited a difference in aneurysm location compared to the control group (0 vs 6.15% localized in the vertebral artery, P = 0.024). No other baseline characteristics showed differences between the cohorts **(Table 1**, **Table 4)**.

**Table 1.** Baseline characteristics of patients administrated with hemostatic agents.

|  | Control cohort<br>(n=65) | Hemostatic<br>medications<br>(n=280) | P value | OR (95% CI) |
| --- | --- | --- | --- | --- |
| Population |  |  |  |  |
| Male | 24 (36.92) | 88 (31.43) | 0.52 | 0.83 (0.48-1.47) |
| Age - y | 61.54 ± 12.29 | 60.48 ± 10.06 | 0.46 | 0.84 (0.52-1.36) |
| Smoking | 6 (9.23) | 27 (9.64) | 0.94 | 1.04 (0.44-2.88) |
| Alcohol | 5 (7.69) | 27 (9.64) | 0.63 | 1.27 (0.51-3.85) |
| Neurosurgical interventions |  |  |  |  |
| Endovascular coiling | 60 (92.31) | 98 (35.00) | <0.001 | 0.14 (0.05-0.32) |
| Neurosurgical clipping | 5 (7.69) | 182 (65.00) |  |  |
| GCS |  |  |  |  |
| 3-8 | 3 (4.62) | 8 (2.86) | 0.75 | 0.79 (0.22-3.76) |
| 9-12 | 6 (9.23) | 23 (8.21) | 0.72 | 1.19 (0.47-3.45) |
| 13-15 | 30 (46.15) | 97 (34.64) | 0.90 | 0.95 (0.39-2.15) |
| Unknown | 26 (40.00) | 152 (54.29) |  |  |
| Complication |  |  |  |  |
| Hypertension | 45 (69.23) | 189 (67.50) | 0.72 | 0.90 (0.49-1.59) |
| Diabetes | 7 (10.77) | 18 (6.43) | 0.25 | 0.57 (0.24-1.53) |
| Coronary heart disease | 4 (6.15) | 9 (3.21) | 0.26 | 0.48 (0.15-1.83) |
| Hyperlipidemia | 1 (1.54) | 1 (0.36) | 0.31 | 0.22 (0.01-5.59) |
| Kidney insufficiency | 1 (1.54) | 3 (1.07) | 0.73 | 0.66 (0.08-13.50) |
| Cardiac insufficiency | 3 (4.62) | 5 (1.79) | 0.19 | 0.36 (0.08-1.78) |
| Intracranial hemorrhage | 6 (9.23) | 16 (5.71) | 0.28 | 0.57 (0.22-1.64) |
| Hydrocephalus | 0 | 6 (2.14) | 0.51 | NA |
| Cerebral ischemia | 10 (15.38) | 48 (17.14) | 0.84 | 1.08 (0.53-2.38) |
| Location |  |  |  |  |
| Anterior Cerebral Artery | 2 (3.08) | 11 (3.93) | 0.69 | 1.34 (0.36-8.78) |
| Middle Cerebral Artery | 13 (20.00) | 46 (16.43) | 0.53 | 0.80 (0.42-1.64) |
| Posterior Cerebral Artery | 2 (3.08) | 3 (1.07) | 0.25 | 0.32 (0.05-2.51) |
| Basilar Artery | 2 (3.08) | 3 (1.07) | 0.25 | 0.32 (0.05-2.51) |
| Internal Carotid Artery | 8 (12.31) | 25 (8.93) | 0.36 | 0.66 (0.30-1.64) |
| Anterior Communicating Artery | 13 (20.00) | 85 (30.36) | 0.06 | 1.83 (0.98-3.66) |
| Posterior Communicating Artery | 20 (30.77) | 96 (34.29) | 0.68 | 1.13 (0.64-2.05) |
| Anterior Choroidal Artery | 1 (1.54) | 2 (0.71) | 0.53 | 0.44 (0.04-9.54) |
| Vertebral Artery | 4 (6.15) | 6 (2.14) | 0.10 | 0.32 (0.09-1.28) |
| **Aneurysm size - mm** | 4.36 (3.30, 6.42) | 4.70 (3.00, 6.00) | 0.72 | 0.91 (0.56-1.50) |
| **Multiple aneurysms** | 17 (26.15) | 63 (22.50) | 0.50 | 0.80 (0.44-1.53) |

Data are presented as n (%), mean ± standard deviation, or median (interquartile range).
NA, odds ratio are not applicable due to the zero value in cohort analysis.
aOR, adjusted odds ratio; CI, confidence interval; GCS, Glasgow Coma Scale; H&H, Hunt and Hess; OR, odds ratio.

Additionally, we observed differences in the timing of administration across different hemostatic regimens. Antifibrinolytics were predominantly given preoperatively, whereas the combination of hemocoagulase and antifibrinolytics was mostly administered both pre- and postoperatively, a detail not previously mentioned in earlier studies **(Supplementary Table 1)**.

### The Impact of Hemostatic Medications on Major Outcomes and Cerebrovascular Events Following aSAH Interventions

Patients with aSAH who received hemostatic medications experienced a delayed treatment initiation compared to the control cohort (adjusted OR [aOR] 4.19, 95% CI 2.48-7.06). However, there were no differences between the two cohorts in terms of LOS (aOR 1.30, 95% CI 0.80-2.12) and mortality rates (aOR 1.69, 95% CI 0.54-7.43) following interventions. Nevertheless, patients perioperatively treated with hemostatic medications demonstrated improved neurological outcomes aOR 0.14, 95% CI 0.02-0.96). Despite this, hemostatic medications did not significantly affect the overall rebleeding rate (aOR 0.35, 95% CI 0.12-1.10) or the incidence of cerebral ischemia (aOR 1.18, 95% CI 0.58-2.56) **(Table 2)**.

**Table 2.**
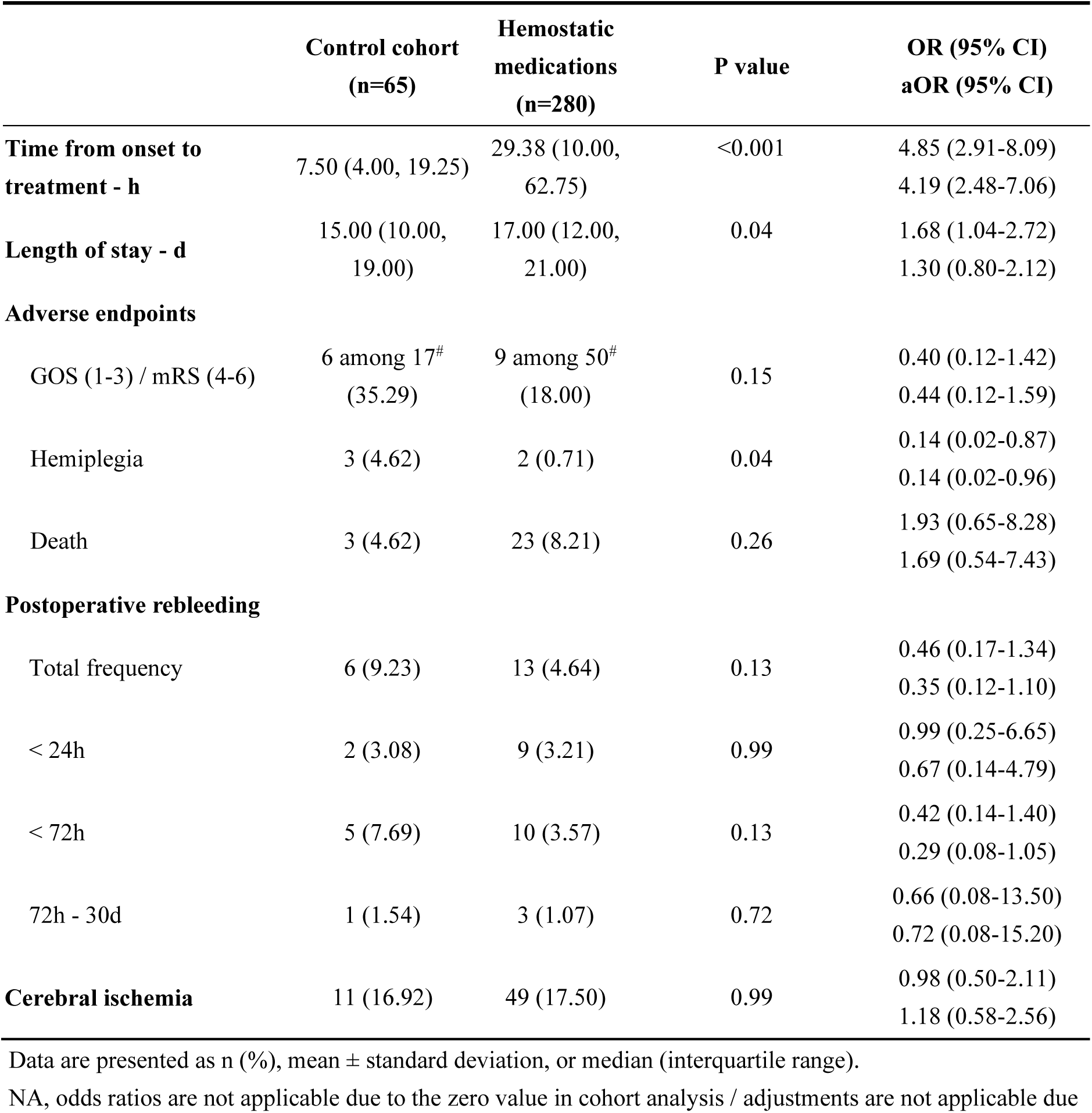
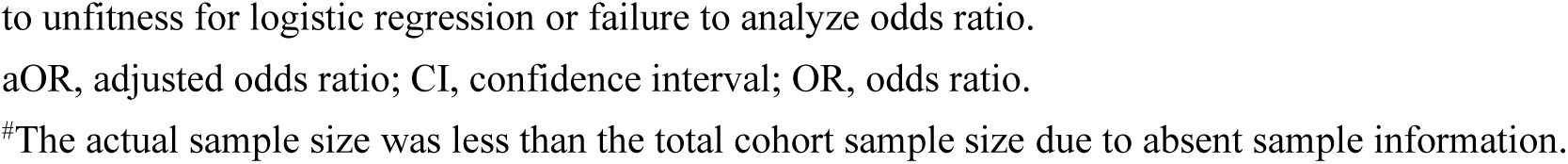
Therapy timing, primary outcomes, and cerebrovascular events in cohorts receiving hemostatic agents.

|  | **Control cohort**<br>**(n=65)** | **Hemostatic**<br>**medications**<br>**(n=280)** | **P value** | **OR (95% CI)**<br>**aOR (95% CI)** |
| --- | --- | --- | --- | --- |
| **Time from onset to**<br>**treatment - h** | 7.50 (4.00, 19.25) | 29.38 (10.00,<br>62.75) | <0.001 | 4.85 (2.91-8.09)<br>4.19 (2.48-7.06) |
| **Length of stay - d** | 15.00 (10.00,<br>19.00) | 17.00 (12.00,<br>21.00) | 0.04 | 1.68 (1.04-2.72)<br>1.30 (0.80-2.12) |
| **Adverse endpoints** |  |  |  |  |
| GOS (1-3) / mRS (4-6) | 6 among 17<sup>#</sup><br>(35.29) | 9 among 50<sup>#</sup><br>(18.00) | 0.15 | 0.40 (0.12-1.42)<br>0.44 (0.12-1.59) |
| Hemiplegia | 3 (4.62) | 2 (0.71) | 0.04 | 0.14 (0.02-0.87)<br>0.14 (0.02-0.96) |
| Death | 3 (4.62) | 23 (8.21) | 0.26 | 1.93 (0.65-8.28)<br>1.69 (0.54-7.43) |
| **Postoperative rebleeding** |  |  |  |  |
| Total frequency | 6 (9.23) | 13 (4.64) | 0.13 | 0.46 (0.17-1.34)<br>0.35 (0.12-1.10) |
| < 24h | 2 (3.08) | 9 (3.21) | 0.99 | 0.99 (0.25-6.65)<br>0.67 (0.14-4.79) |
| < 72h | 5 (7.69) | 10 (3.57) | 0.13 | 0.42 (0.14-1.40)<br>0.29 (0.08-1.05) |
| 72h - 30d | 1 (1.54) | 3 (1.07) | 0.72 | 0.66 (0.08-13.50)<br>0.72 (0.08-15.20) |
| **Cerebral ischemia** | 11 (16.92) | 49 (17.50) | 0.99 | 0.98 (0.50-2.11)<br>1.18 (0.58-2.56) |

Data are presented as n (%), mean ± standard deviation, or median (interquartile range).
NA, odds ratios are not applicable due to the zero value in cohort analysis / adjustments are not applicable due
to unfitness for logistic regression or failure to analyze odds ratio.
aOR, adjusted odds ratio; CI, confidence interval; OR, odds ratio.
<sup>#</sup>The actual sample size was less than the total cohort sample size due to absent sample information.

**Table 3.** Adverse events following the administration of hemostatic agents.

|  | **Control cohort**<br>**(n=65)** | **Hemostatic**<br>**medications**<br>**(n=280)** | **P value** | **OR (95% CI)**<br>**aOR (95% CI)** |
| --- | --- | --- | --- | --- |
| **Intracranial Injuries** |  |  |  |  |
| Increased intracranial pressure | 32 (49.23) | 160 (57.14) | 0.22 | 1.40 (0.82-2.42)<br>0.87 (0.50-1.53) |
| Hydrocephalus | 1 (1.54) | 12 (4.29) | 0.30 | 2.97 (0.58-54.44)<br>2.68 (0.49-49.87) |
| Cerebral vasospasm | 8 (12.31) | 47 (16.79) | 0.29 | 1.54 (0.73-3.66)<br>0.98 (0.44-2.43) |
| Seizures | 0 | 6 (2.14) | 0.60 | NA |
| Brain herniation | 2 (3.08) | 10 (3.57) | 0.89 | 1.11 (0.28-7.36)<br>0.83 (0.19-5.79) |
| Brain edema | 2 (3.08) | 10 (3.57) | 0.89 | 1.11 (0.28-7.36)<br>0.34 (0.06-2.59) |
| Optic nerve injury | 0 | 5 (1.79) | 0.59 | NA |
| Intracranial Infection | 9 (13.85) | 21 (7.50) | 0.10 | 0.48 (0.21-1.16)<br>0.36 (0.15-0.92) |
| **Extracranial Complications** |  |  |  |  |
| Fluid and electrolyte disturbances | 49 (75.38) | 192 (68.57) | 0.33 | 0.74 (0.39-1.34)<br>0.62 (0.32-1.14) |
| Pneumonia | 17 (26.15) | 126 (45.00) | 0.005 | 2.28 (1.76-4.26)<br>1.53 (0.83-2.93) |
| Hyperglycemia | 9 (13.85) | 60 (21.43) | 0.21 | 1.60 (0.78-3.63)<br>1.58 (0.75-3.64) |
| Cardiac insufficiency | 5 (7.69) | 26 (9.29) | 0.76 | 1.17 (0.46-3.56)<br>1.18 (0.45-3.68) |
| Hypoproteinemia | 0 | 13 (4.64) | 0.14 | NA |
| Fever | 1 (1.54) | 7 (2.50) | 0.66 | 1.57 (0.27-29.54)<br>1.49 (0.24-28.87) |
| Organ failure | 1 (1.54) | 7 (2.50) | 0.56 | 1.80 (0.32-33.62)<br>1.83 (0.30-34.90) |
| Shock | 0 | 3 (1.07) | >0.999 | NA |

Data are presented as n (%), mean ± standard deviation, or median (interquartile range).
NA, odds ratios are not applicable due to the zero value in cohort analysis / adjustments are not applicable due to unfitness for logistic regression or failure to analyze odds ratio.
aOR, adjusted odds ratio; CI, confidence interval; OR, odds ratio.
\*P-value versus control group, \*P<0.05, \*\*P<0.01, \*\*\*P<0.001.

### The Impact of Different Combination Regimens of Hemostatic Medications on Major Outcomes and Cerebrovascular Events Following aSAH Interventions

Compared to the cohort not using hemostatic medications, the use of a single class of hemostatic agent significantly improved neurological outcomes post-treatment (aOR 0.11, 95% CI 0.00-0.99). However, combining two (OR 0.18, 95% CI 0.01-1.41) or three (P > 0.999) types of hemostatic agents did not alter the neurological outcomes. There was no statistically significant association between the number of hemostatic agents and patient mortality (Single-agent: aOR 2.01, 95% CI 0.60-9.18; Dual-agent: aOR 1.82, 95% CI 0.35-13.42; Triple-agent: aOR 1.77, 95% CI 0.20-14.29) or LOS (Single-agent: aOR 1.15, 95% CI 0.68-1.94; Dual-agent: aOR 1.64, 95% CI 0.91-2.95; Triple-agent: aOR 1.50, 95% CI 0.58-3.87) **(Table 4** - **5)**.

**Table 4.** Baseline characteristics of patients administrated with different hemostatic agent strategies.

|  | Control cohort<br>(n=65) | Single-agent<br>(n=141) | OR (95% CI) | Dual-agent<br>(n=118) | OR (95% CI) | Triple-agent<br>(n=21) | OR (95% CI) |
| --- | --- | --- | --- | --- | --- | --- | --- |
| **Population** |  |  |  |  |  |  |  |
| Male | 24 (36.92) | 45 (31.91) | 0.80 (0.43-1.49) | 37 (31.36) | 0.78 (0.41-1.48) | 6 (28.57) | 0.68 (0.22-1.93) |
| Age - y | 61.54 ± 12.29 | 60.67 ± 10.13 | 0.87 (0.52-0.38) | 60.16 ± 9.96 | 0.81 (0.47-1.38) | 61.00 ± 10.56 | 0.92 (0.40-2.11) |
| Smoking | 6 (9.23) | 12 (8.51) | 0.91 (0.34-2.74) | 14 (11.86) | 1.32 (0.50-3.90) | 1 (4.76) | 0.49 (0.02-3.12) |
| Alcohol | 5 (7.69) | 15 (10.64) | 1.43 (0.53-4.56) | 11 (9.32) | 1.23 (0.43-4.07) | 1 (4.76) | 0.60 (0.03-4.02) |
| **Neurosurgical interventions** |  |  |  |  |  |  |  |
| Endovascular coiling | 60 (92.31) | 108 (76.60) | 3.67 (1.47-11.15) | 65 (55.08) | 9.78 (3.99-29.54) | 9 (42.86) | 16.00 (4.81-61.37) |
| Neurosurgical clipping | 5 (7.69) | 33 (23.40)** |  | 53 (44.92)**** |  | 12 (57.14)**** |  |
| **GCS** |  |  |  |  |  |  |  |
| 3-8 | 3 (4.62) | 3 (2.13) | 0.51 (0.09-2.86) | 3 (2.54) | 0.90 (0.16-5.13) | 2 (9.52) | 2.67 (0.32-18.57) |
| 9-12 | 6 (9.23) | 14 (9.93) | 1.28 (0.47-3.91) | 8 (6.78) | 1.26 (0.40-4.19) | 1 (4.76) | 0.55 (0.03-3.78) |
| 13-15 | 30 (46.15) | 57 (40.43) | 1.01 (0.39-2.49) | 32 (27.12) | 0.87 (0.31-2.40) | 8 (38.10) | 0.80 (0.18-4.22) |
| Unknown | 26 (40.00) | 67 (47.52) |  | 75 (63.56) |  | 10 (47.62) |  |
| **Complication** |  |  |  |  |  |  |  |
| Hypertension | 45 (69.23) | 99 (70.21) | 1.05 (0.55-1.97) | 78 (66.10) | 0.87 (0.45-1.65) | 12 (57.14) | 0.59 (0.22-1.66) |
| Diabetes | 7 (10.77) | 10 (7.09) | 0.63 (0.23-1.82) | 8 (6.78) | 0.60 (0.21-1.80) | 0 | NA |
| Coronary heart disease | 4 (6.15) | 4 (2.84) | 0.44 (0.10-1.94) | 3 (2.54) | 0.40 (0.08-1.86) | 2 (9.52) | 1.60 (0.21-8.91) |
| Hyperlipidemia | 1 (1.54) | 0 | NA | 1 (0.85) | 0.55 (0.02-13.97) | 0 | NA |
| Kidney insufficiency | 1 (1.54) | 3 (2.13) | 1.39 (0.17-28.41) | 0 | NA | 0 | NA |
| Cardiac insufficiency | 3 (4.62) | 4 (2.84) | 0.60 (0.13-3.14) | 1 (0.85) | 0.18 (0.01-1.41) | 0 | NA |
| Intracranial hemorrhage | 6 (9.23) | 7 (4.96) | 0.51 (0.16-1.66) | 6 (5.08) | 0.53 (0.16-1.75) | 3 (14.29) | 1.64 (0.32-6.90) |
| Hydrocephalus | 0 | 2 (1.42) | NA | 4 (3.39) | NA | 0 | NA |
| Cerebral ischemia | 10 (15.38) | 25 (17.73) | 1.18 (0.54-2.74) | 19 (16.10) | 1.06 (0.47-2.51) | 4 (19.05) | 1.29 (0.32-4.44) |
| **Location** |  |  |  |  |  |  |  |
| Anterior Cerebral Artery | 2 (3.08) | 8 (5.67) | 1.90 (0.46-12.79) | 3 (2.54) | 0.82 (0.13-6.36) | 0 | NA |
| Middle Cerebral Artery | 13 (20.00) | 17 (12.06) | 0.55 (0.25-1.23) | 23 (19.49) | 0.97 (0.46-2.12) | 6 (28.57) | 1.60 (0.49-4.83) |
| Posterior Cerebral Artery | 2 (3.08) | 3 (2.13) | 0.68 (0.11-5.29) | 0 | NA | 0 | NA |
| Basilar Artery | 2 (3.08) | 1 (0.71) | 0.22 (0.01-2.39) | 2 (1.69) | 0.54 (0.06-4.61) | 0 | NA |
| Internal Carotid Artery | 8 (12.31) | 14 (9.93) | 0.78 (0.32-2.06) | 9 (7.63) | 0.59 (0.21-1.64) | 2 (9.52) | 0.75 (0.11-3.32) |
| Anterior Communicating Artery | 13 (20.00) | 43 (30.50) | 1.76 (0.88-3.66) | 38 (32.20) | 1.90 (0.94-4.02) | 4 (19.05) | 0.94 (0.24-3.08) |
| Posterior Communicating Artery | 20 (30.77) | 49 (34.75) | 1.20 (0.64-2.28) | 39 (33.05) | 1.11 (0.58-2.16) | 8 (38.10) | 1.38 (0.48-3.83) |
| Anterior Choroidal Artery | 1 (1.54) | 1 (0.71) | 0.46 (0.02-11.67) | 1 (0.85) | 0.55 (0.02-13.97) | 0 | NA |
| Vertebral Artery | 4 (6.15) | 5 (3.55) | 0.56 (0.14-2.33) | 0* | NA | 1 (4.76) | 0.76 (0.04-5.54) |
| **Aneurysm size - mm** | 4.36 (3.30, 6.42) | 4.40 (3.20, 6.00) | 0.89 (0.51-1.54) | 4.70 (3.00, 6.00) | 0.92 (0.53-1.61) | 5.20 (3.00, 6.20) | 1.04 (0.39-2.80) |
| **Multiple aneurysms** | 17 (26.15) | 34 (24.11) | 0.90 (0.46-1.79) | 24 (20.34) | 0.72 (0.35-1.48) | 5 (23.81) | 0.88 (0.26-2.66) |

Data are presented as n (%), mean ± standard deviation, or median (interquartile range).
NA, odds ratio are not applicable due to the zero value in cohort analysis.
aOR, adjusted odds ratio; CI, confidence interval; GCS, Glasgow Coma Scale; H&H, Hunt and Hess; OR, odds ratio.
\*P-value versus control group, \*P<0.05, \*\*\*\*P<0.0001

**Table 5.**
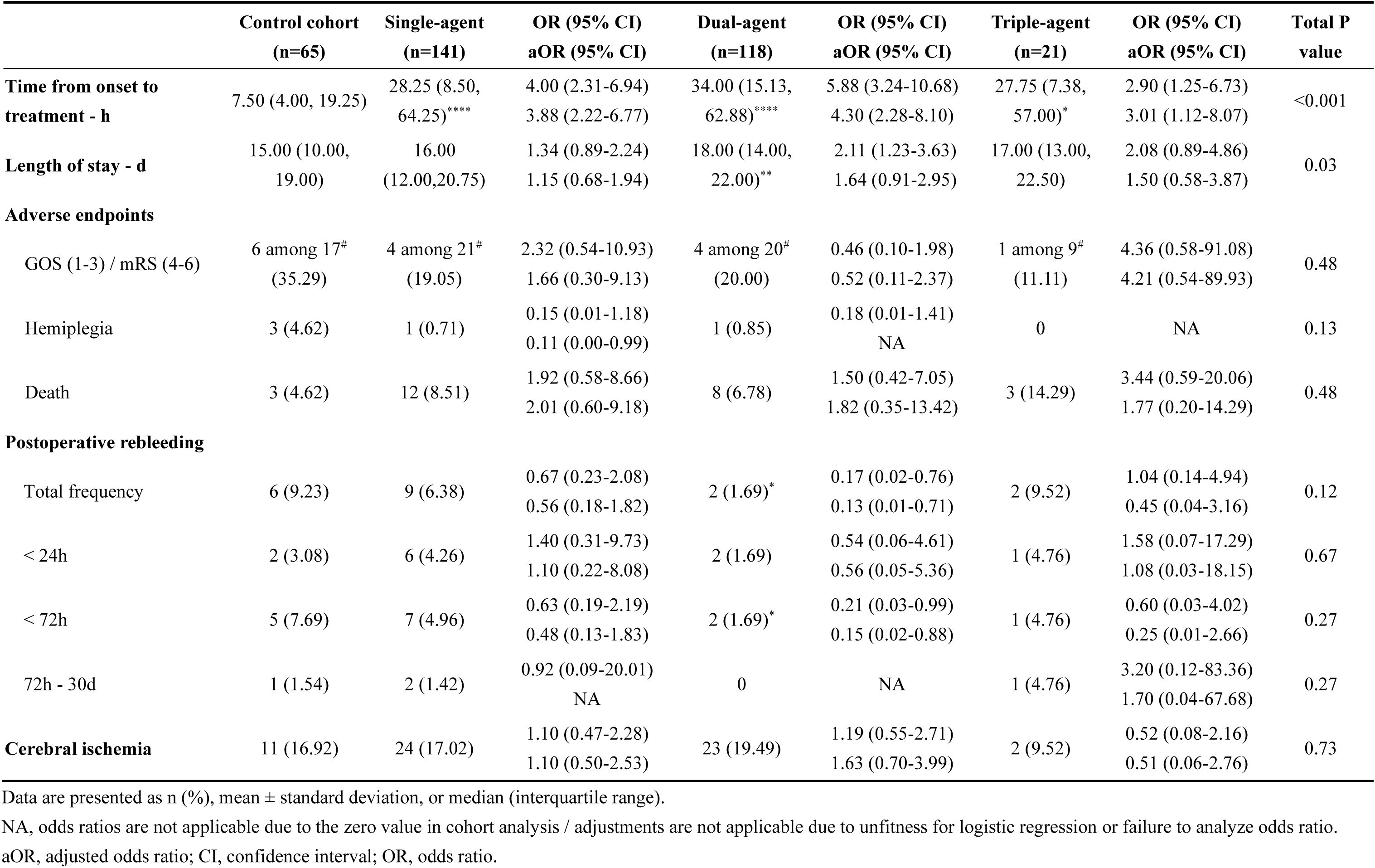
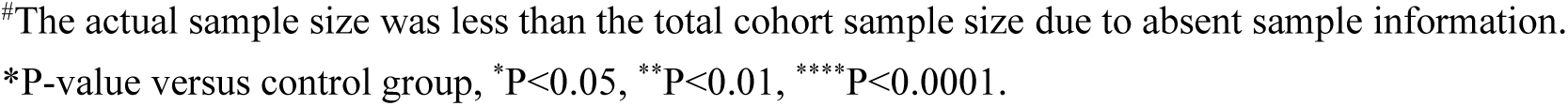
Therapy timing, primary outcomes, and cerebrovascular events across different numbers of combined hemostatic agents.

|  | Control cohort<br>(n=65) | Single-agent<br>(n=141) | OR (95% CI)<br>aOR (95% CI) | Dual-agent<br>(n=118) | OR (95% CI)<br>aOR (95% CI) | Triple-agent<br>(n=21) | OR (95% CI)<br>aOR (95% CI) | Total P<br>value |
| --- | --- | --- | --- | --- | --- | --- | --- | --- |
| **Time from onset to treatment - h** | 7.50 (4.00, 19.25) | 28.25 (8.50, 64.25)**** | 4.00 (2.31-6.94)<br>3.88 (2.22-6.77) | 34.00 (15.13, 62.88)**** | 5.88 (3.24-10.68)<br>4.30 (2.28-8.10) | 27.75 (7.38, 57.00)* | 2.90 (1.25-6.73)<br>3.01 (1.12-8.07) | <0.001 |
| **Length of stay - d** | 15.00 (10.00, 19.00) | 16.00 (12.00,20.75) | 1.34 (0.89-2.24)<br>1.15 (0.68-1.94) | 18.00 (14.00, 22.00)** | 2.11 (1.23-3.63)<br>1.64 (0.91-2.95) | 17.00 (13.00, 22.50) | 2.08 (0.89-4.86)<br>1.50 (0.58-3.87) | 0.03 |
| **Adverse endpoints** |  |  |  |  |  |  |  |  |
| GOS (1-3) / mRS (4-6) | 6 among 17<sup>#</sup><br>(35.29) | 4 among 21<sup>#</sup><br>(19.05) | 2.32 (0.54-10.93)<br>1.66 (0.30-9.13) | 4 among 20<sup>#</sup><br>(20.00) | 0.46 (0.10-1.98)<br>0.52 (0.11-2.37) | 1 among 9<sup>#</sup><br>(11.11) | 4.36 (0.58-91.08)<br>4.21 (0.54-89.93) | 0.48 |
| Hemiplegia | 3 (4.62) | 1 (0.71) | 0.15 (0.01-1.18)<br>0.11 (0.00-0.99) | 1 (0.85) | 0.18 (0.01-1.41)<br>NA | 0 | NA | 0.13 |
| Death | 3 (4.62) | 12 (8.51) | 1.92 (0.58-8.66)<br>2.01 (0.60-9.18) | 8 (6.78) | 1.50 (0.42-7.05)<br>1.82 (0.35-13.42) | 3 (14.29) | 3.44 (0.59-20.06)<br>1.77 (0.20-14.29) | 0.48 |
| **Postoperative rebleeding** |  |  |  |  |  |  |  |  |
| Total frequency | 6 (9.23) | 9 (6.38) | 0.67 (0.23-2.08)<br>0.56 (0.18-1.82) | 2 (1.69)* | 0.17 (0.02-0.76)<br>0.13 (0.01-0.71) | 2 (9.52) | 1.04 (0.14-4.94)<br>0.45 (0.04-3.16) | 0.12 |
| < 24h | 2 (3.08) | 6 (4.26) | 1.40 (0.31-9.73)<br>1.10 (0.22-8.08) | 2 (1.69) | 0.54 (0.06-4.61)<br>0.56 (0.05-5.36) | 1 (4.76) | 1.58 (0.07-17.29)<br>1.08 (0.03-18.15) | 0.67 |
| < 72h | 5 (7.69) | 7 (4.96) | 0.63 (0.19-2.19)<br>0.48 (0.13-1.83) | 2 (1.69)* | 0.21 (0.03-0.99)<br>0.15 (0.02-0.88) | 1 (4.76) | 0.60 (0.03-4.02)<br>0.25 (0.01-2.66) | 0.27 |
| 72h - 30d | 1 (1.54) | 2 (1.42) | 0.92 (0.09-20.01)<br>NA | 0 | NA | 1 (4.76) | 3.20 (0.12-83.36)<br>1.70 (0.04-67.68) | 0.27 |
| **Cerebral ischemia** | 11 (16.92) | 24 (17.02) | 1.10 (0.47-2.28)<br>1.10 (0.50-2.53) | 23 (19.49) | 1.19 (0.55-2.71)<br>1.63 (0.70-3.99) | 2 (9.52) | 0.52 (0.08-2.16)<br>0.51 (0.06-2.76) | 0.73 |

Data are presented as n (%), mean ± standard deviation, or median (interquartile range).
NA, odds ratios are not applicable due to the zero value in cohort analysis / adjustments are not applicable due to unfitness for logistic regression or failure to analyze odds ratio.
aOR, adjusted odds ratio; CI, confidence interval; OR, odds ratio.

**Table 6.**
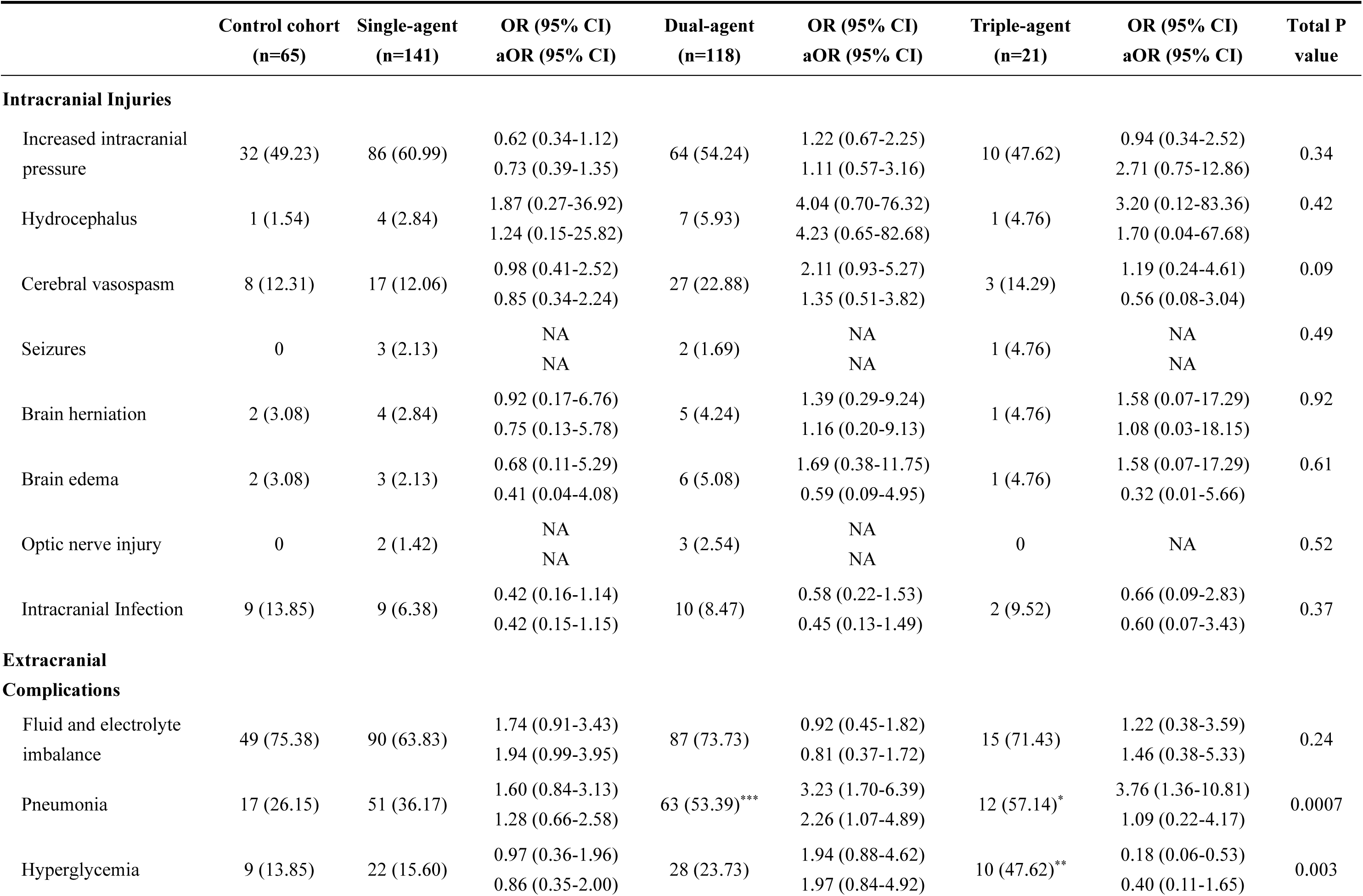
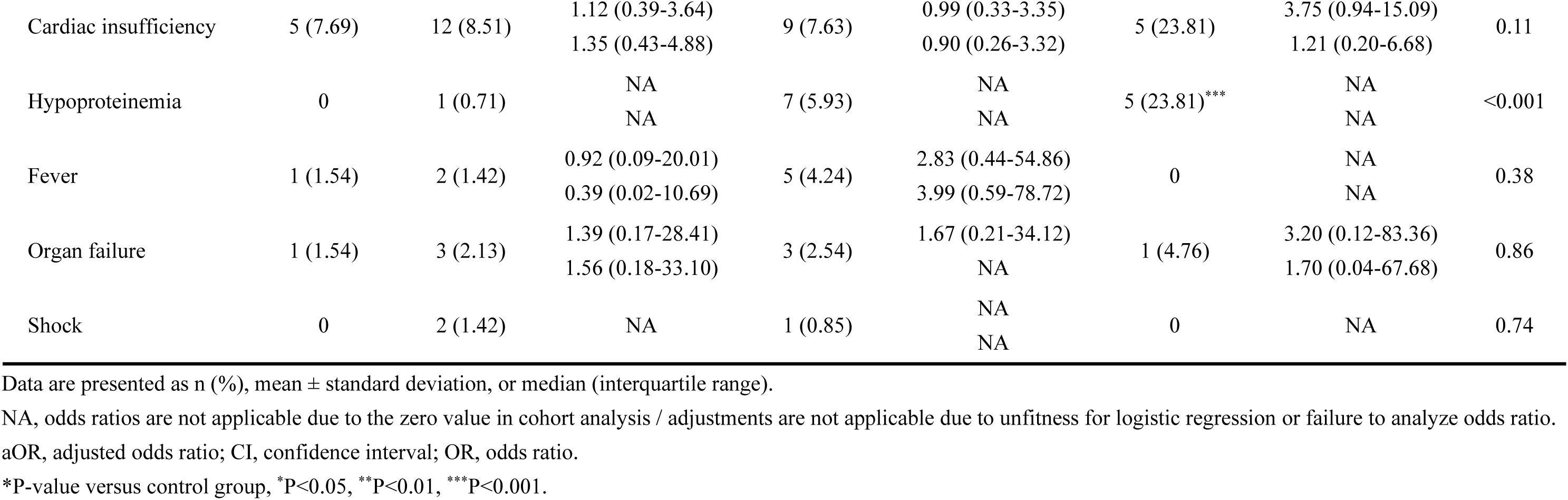
Adverse events following the administration of different numbers of combined hemostatic agents.

Compared to the control cohort, the combination of two hemostatic agents demonstrated a significant reduction in both the overall rebleeding rate (aOR 0.13, 95% CI 0.01-0.71) and the rebleeding rate within 72 hours post-treatment in aSAH patients (aOR 0.15, 95% CI 0.02-0.88). However, no significant differences in rebleeding events were observed between the cohorts using either a single hemostatic agent (aOR 0.56, 95% CI 0.18-1.82) or three agents (aOR 0.45, 0.04-3.16) compared to the control cohort. Additionally, none of the hemostatic medication regimens, whether single (aOR 1.10, 95% CI 0.50-2.53), double (aOR 1.63, 95% CI 0.70-3.99), or triple (aOR 0.51, 95% CI 0.06-2.76) combinations, showed a significant impact on the incidence of cerebral ischemia following neurosurgical interventions relative to the control cohort **(Table 4** - **5)**.

Further analysis was conducted to evaluate the impact of different hemostatic medication regimens on major outcomes and cerebrovascular events in aSAH patients following invasive treatments. Compared to the control cohort, the use of hemocoagulase alone (Poor GOS/mRS: aOR 2.83, 95% CI 0.46-24.48; Hemiplegia: aOR 0.28, 95% CI 0.01-2.65; Mortality: OR 0.75, 95% CI 0.10-4.69) or antifibrinolytic agents alone (Poor GOS/mRS: P > 0.999; Hemiplegia: P = 0.16; Mortality: aOR 3.05, 95% CI 0.84-15.64) did not improve adverse endpoints. Similarly, the combination of hemocoagulase with a single antifibrinolytic agent (Poor GOS/mRS: aOR 0.39, 95% CI 0.07-1.91; Hemiplegia: OR 0.18, 95% CI 0.01-1.48; Mortality: aOR 1.68, 95% CI 0.33-12.35), the combination of two antifibrinolytic agents (Poor GOS/mRS: P = 0.89; Hemiplegia: P > 0.999; Mortality: OR 5,17, 95% CI 0.23-53.19) and the combination of hemocoagulase with two antifibrinolytic agents (Poor GOS/mRS: aOR 4.21, 95% CI 0.54-89.93; Hemiplegia: P > 0.999; Mortality: aOR 1.77, 95% CI 0.20-14.29) did not show significant improvements. Additionally, no significant differences were noted across the different regimens regarding LOS (Hemocoagulase-only: aOR 1.01, 95% CI 0.46-2.18; Antifibrinolytic-only: aOR 1.25, 95% CI 0.70-2.24; Hemocoagulase and single antifibrinolytic: aOR 1.65, 95% CI 0.91-2.99; Double antifibrinolytics: OR 3.56, 95% CI 0.54-23.42; Hemocoagulase and double antifibrinolytic: aOR 1.50, 95% CI 0.58-3.87).

However, the combination of hemocoagulase with a single antifibrinolytic agent demonstrated a significant reduction in both the overall rebleeding rate (aOR 0.13, 95% CI 0.02-0.74) and the rebleeding rate within 72 hours post-intervention (aOR 0.15, 95% CI 0.02-0.92). In contrast, no significant differences in rebleeding rates were observed for other regimens compared to those who did not use hemostatic agents (Hemocoagulase-only: aOR 0.16, 95% CI 0.01-1.00; Antifibrinolytic-only: aOR 1.28, 95% CI 0.37-4.85; Double antifibrinolytics: P > 0.999; Hemocoagulase and double antifibrinolytics: aOR 0.45, 95% CI 0.04-3.16) **(Table 7 - 8)**.

**Table 7.** Baseline characteristics of cohorts by strategies for the administration of hemostatic agents.

|  | Control<br>cohort<br>(n=65) | Hemocoagula<br>se-only<br>(n=57) | OR (95% CI) | Antifibrino<br>lytic-only<br>(n=84) | OR (95% CI) | Hemocoagul<br>ase & single<br>antifibrinoly<br>tic (n=113) | OR (95% CI) | Double<br>antifibrinol<br>ytics (n=5) | OR (95% CI) |
| --- | --- | --- | --- | --- | --- | --- | --- | --- | --- |
| Population |  |  |  |  |  |  |  |  |  |
| Male | 24 (36.92) | 21 (36.84) | 0.99 (0.47-2.08) | 24 (28.57) | 0.68 (0.34-1.36) | 34 (30.09) | 0.74 (0.39-1.41) | 3 (60.00) | 2.56 (0.40-20.51) |
| Age - y | 61.54 ±<br>12.29 | 58.12 ± 8.92 | 0.59 (0.32-1.10) | 62.39 ±<br>10.59 | 1.13 (0.64-2.00) | 60.27 ±<br>10.07 | 0.82 (0.48-1.40) | 57.60 ± 7.44 | 0.62 (0.15-2.61) |
| Smoking | 6 (9.23) | 8 (14.04) | 1.60 (0.52-5.17) | 4 (4.76) | 0.49 (0.12-1.80) | 14 (12.39) | 1.39 (0.53-4.10) | 0 | NA |
| Alcohol | 5 (7.69) | 8 (14.04) | 1.96 (0.61-6.84) | 7 (8.33) | 1.09 (0.33-3.84) | 10 (8.85) | 1.16 (0.39-3.89) | 1 (20.00) | 3.00 (0.14-25.98) |
| Neurosurgical<br>interventions |  |  |  |  |  |  |  |  |  |
| Endovascular coiling | 60 (92.31) | 39 (68.42) | 5.54 (2.02-17.89) | 69 (82.14) | 2.61 (0.95-8.40) | 62 (54.87) | 9.87 (4.01-29.86) | 3 (60.00) | 8.00 (0.90-61.08) |
| Neurosurgical clipping | 5 (7.69) | 18 (31.58)*** |  | 15 (17.86) |  | 51(45.13)**** |  | 2 (40.00) |  |
| GCS |  |  |  |  |  |  |  |  |  |
| 3-8 | 3 (4.62) | 0 | NA | 3 (3.57) | 0.95 (0.16-5.41) | 2 (1.77) | 0.60 (0.08-3.81) | 1 (100.00) | NA |
| 9-12 | 6 (9.23) | 9 (15.79) | 2.06 (0.66-6.90) | 5 (5.95) | 0.76 (0.20-2.77) | 8 (7.08) | 1.29 (0.41-4.32) | 0 | NA |
| 13-15 | 30 (46.15) | 24 (42.11) | 0.80 (0.27-2.35) | 33 (39.29) | 1.24 (0.42-3.70) | 32 (28.32) | 0.96 (0.34-2.70) | 0 | NA |
| Unknown | 26 (40.00) | 24 (42.11) |  | 43 (51.19) |  | 71 (62.83) |  | 4 |  |
| Complication |  |  |  |  |  |  |  |  |  |
| Hypertension | 45 (69.23) | 35 (61.40) | 0.71 (0.33-1.50) | 64 (76.19) | 1.42 (0.68-2.96) | 76 (67.26) | 0.91 (0.47-1.75) | 2 (40.00) | 0.30 (0.04-1.92) |
| Diabetes | 7 (10.77) | 2 (3.51) | 0.30 (0.04-1.31) | 8 (9.52) | 0.87 (0.30-2.62) | 8 (7.08) | 0.63 (0.22-1.88) | 0 | NA |
| Coronary heart disease | 4 (6.15) | 2 (3.51) | 0.55 (0.07-2.96) | 2 (2.38) | 0.37 (0.05-1.97) | 3 (2.65) | 0.42 (0.08-1.94) | 0 | NA |
| Hyperlipidemia | 1 (1.54) | 0 | NA | 0 | NA | 1 (0.88) | 0.57 (0.02-14.60) | 0 | NA |
| Kidney insufficiency | 1 (1.54) | 0 | NA | 3 (3.57) | 2.37 (0.30-48.53) | 0 | NA | 0 | NA |
| Cardiac insufficiency | 3 (4.62) | 2 (3.51) | 0.75 (0.10-4.69) | 2 (2.38) | 0.50 (0.06-3.13) | 1 (0.88) | 0.18 (0.01-1.48) | 0 | NA |
| Intracranial hemorrhage | 6 (9.23) | 6 (10.52) | 1.16 (0.34-3.91) | 1 (1.19)* | 0.12 (0.01-0.72) | 6 (5.31) | 0.55 (0.16-1.84) | 0 | NA |
| Hydrocephalus | 0 | 0 | NA | 2 (2.38) | NA | 3 (2.65) | NA | 1 (20.00) | NA |
| Cerebral ischemia | 10 (15.38) | 18 (31.58)* | 2.54 (1.08-6.28) | 7 (8.33) | 0.50 (0.17-1.38) | 18 (15.93) | 1.04 (0.46-2.50) | 1 (20.00) | 1.38 (0.07-10.58) |
| **Location** |  |  |  |  |  |  |  |  |  |
| Anterior Cerebral Artery | 2 (3.08) | 5 (8.77) | 3.03 (0.62-21.75) | 3 (3.57) | 1.17 (0.19-9.06) | 3 (2.65) | 0.86 (0.14-6.65) | 0 | NA |
| Middle Cerebral Artery | 13 (20.00) | 10 (17.54) | 0.85 (0.33-2.12) | 7 (8.33)* | 0.36 (0.13-0.95) | 23 (20.35) | 1.02 (0.48-2.24) | 0 | NA |
| Posterior Cerebral Artery | 2 (3.08) | 1 (1.75) | 0.56 (0.02-6.02) | 2 (2.38) | 0.77 (0.09-6.54) | 0 | NA | 0 | NA |
| Basilar Artery | 2 (3.08) | 0 | NA | 1 (1.19) | 0.38 (0.02-4.04) | 2 (1.77) | 0.57 (0.07-4.82) | 0 | NA |
| Internal Carotid Artery | 8 (12.31) | 5 (8.77) | 0.68 (0.20-2.19) | 9 (10.71) | 0.86 (0.31-2.41) | 9 (7.96) | 0.62 (0.22-1.72) | 0 | NA |
| Anterior Communicating Artery | 13 (20.00) | 18 (31.58) | 1.85 (0.81-4.28) | 25 (29.76) | 1.70 (0.80-3.73) | 36 (31.86) | 1.87 (0.92-3.97) | 2 (40.00) | 2.67 (0.33-17.75) |
| Posterior Communicating Artery | 20 (30.77) | 15 (26.32) | 0.80 (0.36-1.77) | 34 (40.48) | 1.53 (0.78-3.06) | 37 (32.74) | 1.10 (0.57-2.14) | 2 (40.00) | 1.50 (0.19-9.73) |
| Anterior Choroidal Artery | 1 (1.54) | 1 (1.75) | 1.14 (0.04-29.32) | 0 | NA | 1 (0.88) | 0.57 (0.02-14.60) | 0 | NA |
| Vertebral Artery | 4 (6.15) | 2 (3.51) | 0.55 (0.07-2.96) | 3 (3.57) | 0.56 (0.11-2.65) | 0* | NA | 0 | NA |
| **Aneurysm size - mm** | 4.36 (3.30, 6.42) | 3.60 (2.70, 5.29)* | 0.48 (0.24-0.96) | 5.00 (3.36, 6.50) | 1.27 (0.70-2.30) | 4.70 (3.00, 6.00) | 0.90 (0.52-1.59) | 5.30 (3.40, 6.68) | 1.38 (0.26-7.28) |
| **Multiple aneurysms** | 17 (26.15) | 14 (24.56) | 0.92 (0.40-2.08) | 20 (23.81) | 0.88 (0.42-1.88) | 23 (20.35) | 0.72 (0.35-1.49) | 1 (20.00) | 0.70 (0.03-5.20) |

Data are presented as n (%), mean ± standard deviation, or median (interquartile range).
NA, odds ratio are not applicable due to the zero value in cohort analysis.
aOR, adjusted odds ratio; CI, confidence interval; GCS, Glasgow Coma Scale; H&H, Hunt and Hess; OR, odds ratio.
\*P-value versus control group, \*P<0.05, \*\*\*P<0.001, \*\*\*\*P<0.0001.

**Table 8.**
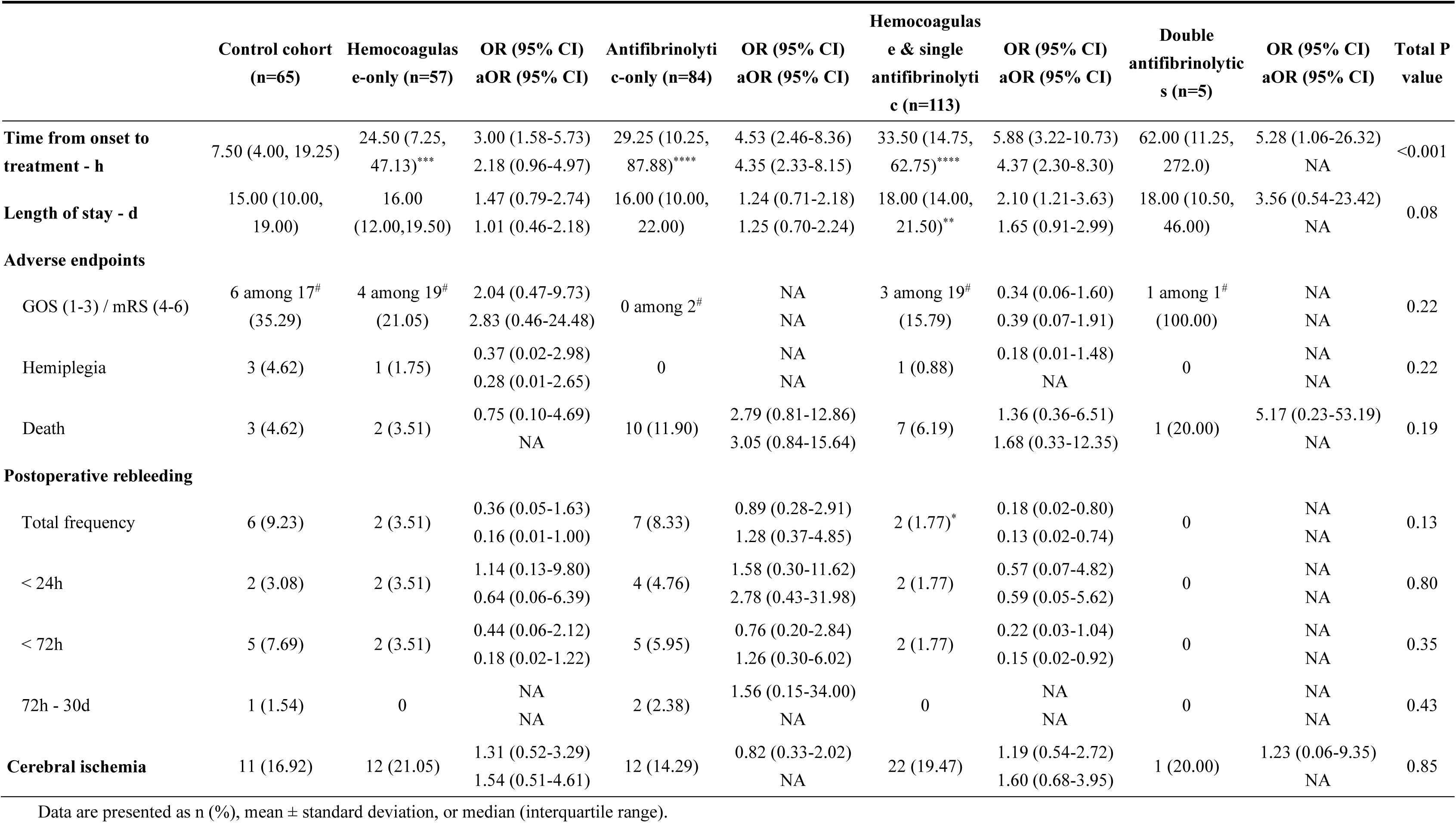
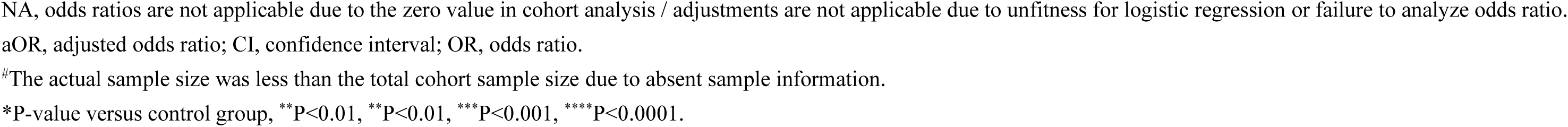
Therapy timing, primary outcomes, and cerebrovascular events by strategies for the administration of hemostatic agents.

|  | Control cohort<br>(n=65) | Hemocoagulas<br>e-only (n=57) | OR (95% CI)<br>aOR (95% CI) | Antifibrinolyti<br>c-only (n=84) | OR (95% CI)<br>aOR (95% CI) | Hemocoagulas<br>e & single<br>antifibrinolyti<br>c (n=113) | OR (95% CI)<br>aOR (95% CI) | Double<br>antifibrinolytic<br>s (n=5) | OR (95% CI)<br>aOR (95% CI) | Total P<br>value |
| --- | --- | --- | --- | --- | --- | --- | --- | --- | --- | --- |
| **Time from onset to**<br>**treatment - h** | 7.50 (4.00, 19.25) | 24.50 (7.25,<br>47.13)*** | 3.00 (1.58-5.73)<br>2.18 (0.96-4.97) | 29.25 (10.25,<br>87.88)**** | 4.53 (2.46-8.36)<br>4.35 (2.33-8.15) | 33.50 (14.75,<br>62.75)**** | 5.88 (3.22-10.73)<br>4.37 (2.30-8.30) | 62.00 (11.25,<br>272.0) | 5.28 (1.06-26.32)<br>NA | <0.001 |
| **Length of stay - d** | 15.00 (10.00,<br>19.00) | 16.00<br>(12.00,19.50) | 1.47 (0.79-2.74)<br>1.01 (0.46-2.18) | 16.00 (10.00,<br>22.00) | 1.24 (0.71-2.18)<br>1.25 (0.70-2.24) | 18.00 (14.00,<br>21.50)** | 2.10 (1.21-3.63)<br>1.65 (0.91-2.99) | 18.00 (10.50,<br>46.00) | 3.56 (0.54-23.42)<br>NA | 0.08 |
| **Adverse endpoints** |  |  |  |  |  |  |  |  |  |  |
| GOS (1-3) / mRS (4-6) | 6 among 17<sup>#</sup><br>(35.29) | 4 among 19<sup>#</sup><br>(21.05) | 2.04 (0.47-9.73)<br>2.83 (0.46-24.48) | 0 among 2<sup>#</sup> | NA<br>NA | 3 among 19<sup>#</sup><br>(15.79) | 0.34 (0.06-1.60)<br>0.39 (0.07-1.91) | 1 among 1<sup>#</sup><br>(100.00) | NA<br>NA | 0.22 |
| Hemiplegia | 3 (4.62) | 1 (1.75) | 0.37 (0.02-2.98)<br>0.28 (0.01-2.65) | 0 | NA<br>NA | 1 (0.88) | 0.18 (0.01-1.48)<br>NA | 0 | NA<br>NA | 0.22 |
| Death | 3 (4.62) | 2 (3.51) | 0.75 (0.10-4.69)<br>NA | 10 (11.90) | 2.79 (0.81-12.86)<br>3.05 (0.84-15.64) | 7 (6.19) | 1.36 (0.36-6.51)<br>1.68 (0.33-12.35) | 1 (20.00) | 5.17 (0.23-53.19)<br>NA | 0.19 |
| **Postoperative rebleeding** |  |  |  |  |  |  |  |  |  |  |
| Total frequency | 6 (9.23) | 2 (3.51) | 0.36 (0.05-1.63)<br>0.16 (0.01-1.00) | 7 (8.33) | 0.89 (0.28-2.91)<br>1.28 (0.37-4.85) | 2 (1.77)* | 0.18 (0.02-0.80)<br>0.13 (0.02-0.74) | 0 | NA<br>NA | 0.13 |
| < 24h | 2 (3.08) | 2 (3.51) | 1.14 (0.13-9.80)<br>0.64 (0.06-6.39) | 4 (4.76) | 1.58 (0.30-11.62)<br>2.78 (0.43-31.98) | 2 (1.77) | 0.57 (0.07-4.82)<br>0.59 (0.05-5.62) | 0 | NA<br>NA | 0.80 |
| < 72h | 5 (7.69) | 2 (3.51) | 0.44 (0.06-2.12)<br>0.18 (0.02-1.22) | 5 (5.95) | 0.76 (0.20-2.84)<br>1.26 (0.30-6.02) | 2 (1.77) | 0.22 (0.03-1.04)<br>0.15 (0.02-0.92) | 0 | NA<br>NA | 0.35 |
| 72h - 30d | 1 (1.54) | 0 | NA<br>NA | 2 (2.38) | 1.56 (0.15-34.00)<br>NA | 0 | NA<br>NA | 0 | NA<br>NA | 0.43 |
| **Cerebral ischemia** | 11 (16.92) | 12 (21.05) | 1.31 (0.52-3.29)<br>1.54 (0.51-4.61) | 12 (14.29) | 0.82 (0.33-2.02)<br>NA | 22 (19.47) | 1.19 (0.54-2.72)<br>1.60 (0.68-3.95) | 1 (20.00) | 1.23 (0.06-9.35)<br>NA | 0.85 |

Data are presented as n (%), mean ± standard deviation, or median (interquartile range).
NA, odds ratios are not applicable due to the zero value in cohort analysis / adjustments are not applicable due to unfitness for logistic regression or failure to analyze odds ratio.
aOR, adjusted odds ratio; CI, confidence interval; OR, odds ratio.

### The Impact of Hemostatic Medications on Other Adverse Events Following aSAH Interventions

Compared to control cohort, aSAH patients who received hemostatic agents exhibited a significantly lower incidence of intracranial infection following neurosurgical interventions (aOR 0.36, 95% CI 0.15-0.92) **(Table 3)**. However, aSAH patients treated with two hemostatic agents showed an increased rate of pulmonary infections (aOR 2.26, 95% CI 1.07-4.89) **(Table 6)**. A similar trend was observed in patients receiving the combination of hemocoagulase and a single antifibrinolytic agent (aOR 2.25, 95% CI 1.08-4.84). Furthermore, patients using hemocoagulase alone demonstrated significantly lower rates of IICP (aOR 0.42, 95% CI 0.18-0.94) and hyperglycemia (aOR 0.25, 95% CI 0.06-0.91) compared to the control cohort. Conversely, the incidence of fluid and electrolyte disturbances was higher in patients receiving hemocoagulase monotherapy (aOR 3.19, 95% CI 1.22-8.75) **(Table 9)**.

**Table 9.** Adverse events following the administration of different strategies for hemostatic agents.

|  | Control<br>cohort (n=65) | Hemocoagula<br>se-only<br>(n=57) | OR (95% CI)<br>aOR (95% CI) | Antifibrinolytic-only (n=84) | OR (95% CI)<br>aOR (95% CI) | Hemocoagulase<br>& single<br>antifibrinolytic<br>(n=113) | OR (95% CI)<br>aOR (95% CI) | double<br>antifibrinolytics<br>(n=5) | OR (95% CI)<br>aOR (95% CI) | Total P<br>value |
| --- | --- | --- | --- | --- | --- | --- | --- | --- | --- | --- |
| **Intracranial Injuries** |  |  |  |  |  |  |  |  |  |  |
| Increased intracranial pressure | 32 (49.23) | 42 (73.68)** | 0.35 (0.16-0.73)<br>0.42 (0.18-0.94) | 44 (52.38) | 0.88 (0.46-1.69)<br>0.79 (0.40-1.55) | 61 (53.98) | 1.21 (0.66-2.23)<br>1.16 (0.59-2.29) | 3 (60.00) | 1.55 (0.24-12.33)<br>NA | 0.06 |
| Hydrocephalus | 1 (1.54) | 2 (3.51) | 2.33 (0.22-50.84)<br>NA | 2 (2.38) | 1.56 (0.15-34.00)<br>NA | 6 (5.31) | 3.59 (0.59-68.56)<br>3.39 (0.48-67.85) | 1 (20.00) | 16.00 (0.56-461.2)<br>NA | 0.21 |
| Cerebral vasospasm | 8 (12.31) | 12 (21.05) | 1.90 (0.72-5.23)<br>1.90 (0.67-5.61) | 5 (5.95) | 0.45 (0.13-1.42)<br>NA | 26 (23.01) | 2.13 (0.94-5.33)<br>1.28 (0.48-3.66) | 1 (20.00) | 1.78 (0.08-14.10)<br>NA | 0.02 |
| Seizures | 0 | 3 (5.26) | NA<br>NA | 0 | NA<br>NA | 2 (1.77) | NA<br>NA | 0 | NA<br>NA | 0.11 |
| Brain herniation | 2 (3.08) | 1 (1.75) | 0.56 (0.02-6.02)<br>0.39 (0.02-4.68) | 3 (3.57) | 1.17 (0.19-9.06)<br>NA | 5 (4.42) | 1.46 (0.30-10.39)<br>1.22 (0.21-9.58) | 0 | NA<br>NA | 0.90 |
| Brain edema | 2 (3.08) | 3 (5.26) | 1.75 (0.28-13.65)<br>0.45 (0.04-3.74) | 0 | NA<br>NA | 6 (5.31) | 1.77 (0.39-12.30)<br>0.62 (0.10-5.18) | 0 | NA<br>NA | 0.29 |
| Optic nerve injury | 0 | 1 (1.75) | NA<br>NA | 1 (1.19) | NA<br>NA | 3 (2.65) | NA<br>NA | 0 | NA<br>NA | 0.72 |
| Intracranial Infection | 9 (13.85) | 2 (3.51)* | 0.23 (0.03-0.93)<br>0.24 (0.03-1.06) | 7 (8.33) | 0.56 (0.19-1.61)<br>NA | 10 (8.85) | 0.60 (0.23-1.60)<br>0.48 (0.14-1.57) | 0 | NA<br>NA | 0.33 |
| **Extracranial Complications** |  |  |  |  |  |  |  |  |  |  |
| Fluid and electrolyte disturbances | 49 (75.38) | 35 (61.40) | 1.92 (0.89-4.24)<br>3.19 (1.22-8.75) | 55 (65.48) | 1.62 (0.79-3.38)<br>1.92 (0.90-4.22) | 82 (72.57) | 0.86 (0.42-1.72)<br>0.76 (0.35-1.62) | 5 (100.00) | NA<br>NA | 0.19 |
| Pneumonia | 17 (26.15) | 28 (49.12)** | 2.73 (1.29-5.91)<br>1.80 (0.67-4.82) | 23 (27.38) | 1.06 (0.51-2.24)<br>1.06 (0.50-2.30) | 62 (54.87)*** | 3.43 (1.79-6.82)<br>2.25 (1.08-4.84) | 1 (20.00) | 0.70 (0.03-5.20)<br>NA | <0.001 |
| Hyperglycemia | 9 (13.85) | 17 (29.82)* | 0.38 (0.15-0.92)<br>0.25 (0.06-0.91) | 5 (5.95) | 2.54 (0.83-8.64)<br>2.01 (0.60-7.19) | 27 (23.89) | 1.95 (0.88-4.68)<br>1.94 (0.82-4.88) | 1 (20.00) | 1.56 (0.08-12.12)<br>NA | 0.002 |
| Cardiac insufficiency | 5 (7.69) | 6 (10.53) | 1.41 (0.40-5.16)<br>1.80 (0.26-16.41) | 6 (7.14) | 0.92 (0.26-3.34)<br>1.22 (0.32-5.20) | 9 (7.96) | 1.04 (0.34-3.51)<br>0.95 (0.27-3.49) | 0 | NA<br>NA | 0.91 |
| Hypoproteinemia | 0 | 0 | NA<br>NA | 1 (1.19) | NA<br>NA | 7 (6.19) | NA<br>NA | 0 | NA<br>NA | 0.04 |
| Fever | 1 (1.54) | 2 (3.51) | 2.33 (0.22-50.84)<br>1.08 (0.08-26.98) | 0 | NA<br>NA | 5 (4.42) | 2.96 (0.46-57.41)<br>4.19 (0.62-82.81) | 0 | NA<br>NA | 0.34 |
| Organ failure | 1 (1.54) | 0 | 2.37 (0.30-48.53)<br>NA | 3 (3.57) | 2.37 (0.30-48.53)<br>NA | 3 (2.65) | 1.74 (0.22-35.68)<br>NA | 0 | NA<br>NA | 0.66 |
| Shock | 0 | 0 | NA<br>NA | 2 (2.38) | NA<br>NA | 1 (0.88) | NA<br>NA | 0 | NA<br>NA | 0.54 |

Data are presented as n (%), mean ± standard deviation, or median (interquartile range).
NA, odds ratios are not applicable due to the zero value in cohort analysis / adjustments are not applicable due to unfitness for logistic regression or failure to analyze odds ratio.
aOR, adjusted odds ratio; CI, confidence interval; OR, odds ratio.
\*P-value versus control group, \*P<0.05, \*\*P<0.01, \*\*\*P<0.001.

## Discussion

Rebleeding within the first 24 hours following aSAH escalates the risk of adverse outcomes, and timely interventions during this critical period can markedly improve patient prognosis[26]. Nevertheless, the risk of rebleeding remains even after neurosurgical interventions, which is closely associated with heightened rate of adverse outcomes[27,28]. Clinically, hemostatic medications are primarily used in aSAH to delay rebleeding, thereby affording more time for definitive therapeutic measures. Our retrospective analysis, encompassing a decade of data from three medical institutions, focused on aSAH patients undergoing endovascular therapy or microsurgical clipping. The findings revealed that 81.16% of patients received hemostatic medications perioperatively, predominantly antifibrinolytics and hemocoagulase. The data also indicate that among aSAH patients who received hemostatic medications, the timing of vascular repair treatment was somewhat delayed, suggesting these medications might be used to provide additional time for patients unable to immediately undergo neurosurgical interventions, although their efficacy remains a topic of debate[29,30]. Moreover, nearly half (49.64%) of patients were treated with multiple hemostatic regimens, a practice that has not been previously investigated. Consequently, our study aims to elucidate the impact of different hemostatic strategies on clinical outcomes, including rebleeding, cerebral ischemia, and other adverse events, following aSAH interventions. Through adjusting for baseline confounders, we aspire to identify potential strategies to control rebleeding risks and improve outcomes, thereby providing evidence-based guidance for the management of aSAH.

### Short-term use of hemostatic medications does not increase ischemia risk following aSAH interventions but improves neurological outcomes

Currently, the increased risk of thromboembolism associated with antifibrinolytics has sparked significant controversy regarding their clinical benefits[13,15,31]. Studies have shown that while long-term use of antifibrinolytics can effectively reduce rebleeding, it increases the risk of cerebral ischemia. In contrast, ultra-short-term use does not elevate the risk of cerebral ischemia and also fails to reduce the incidence of rebleeding. Furthermore, multiple studies suggest no substantial improvement in patient functional outcomes[5,32,33]. In our study, the median duration for antifibrinolytic medications was only 1.5 days (IQR 1.00, 2.00), and this brief period did not increase the risk of cerebral ischemia or notably impact clinical outcomes, consistent with previous findings[11,14,16,33].

Hemocoagulase, another commonly utilized hemostatic agent, is noted for its lower propensity to induce thromboembolism[18]. Although primarily employed for superficial local coagulation during surgeries, there is limited evidence supporting its efficacy in aSAH treatment. Despite this, over the 10-year period covered by our study, up to 68.21% of aSAH patients received short-term hemocoagulase treatment during perioperative period of neurosurgical interventions. Our analysis revealed that short-term use of hemocoagulase, either alone or in combination with antifibrinolytics, had no significant effect on the occurrence of cerebral ischemia or patient outcomes. However, across all evaluated hemostatic regimens, besides observing no change in cerebral ischemia incidence, we noted a marked reduction in hemiplegia rates, suggesting potential neuroprotective benefits of short-term hemostatic medications during the perioperative period of aSAH.

### Combination Administration with hemocoagulase and a single antifibrinolytic agent reduces rebleeding incidence following aSAH interventions

In clinical practice, thrombins and antifibrinolytics are frequently used as adjuvants during endovascular treatment and clipping to prevent rebleeding in aSAH patients. Our study revealed that 50.36% of aSAH patients received monotherapy with a single hemostatic agent, while the remaining 49.64% were treated with combination regimens involving two or more hemostatic agents. Among those receiving monotherapy, 40.42% were administered hemocoagulase, whereas 59.58% received antifibrinolytics, including tranexamic acid, aminomethylbenzoic acid, or epsilon-aminocaproic acid. The use of antifibrinolytics alone for preventing rebleeding in aSAH remains controversial[14,15,33], with limited systematic research on their role post-intervention. Similarly, there is insufficient evidence supporting the efficacy of hemocoagulase in aSAH treatment. Our findings indicate that monotherapy with either hemocoagulase or antifibrinolytics did not significantly reduce perioperative rebleeding rates compared to the control cohort, suggesting that single-agent hemostatic regimens may not effectively decrease post-treatment rebleeding in aSAH patients.

Given that nearly half of the aSAH patients received combinations of two or more hemostatic drugs perioperatively, we explored the impact of these regimens on rebleeding post-intervention. The majority of patients who received dual-agent combinations experienced significant reductions in overall rebleeding and rebleeding within 72 hours following neurosurgical interventions. Notably, the most commonly used regimen involved combining hemocoagulase with a single antifibrinolytic agent, which significantly reduced both overall and early rebleeding risks. This finding is particularly significant in the lack of current evidence-based recommendations. However, combinations of two antifibrinolytics or three hemostatic agents did not demonstrate any advantage over the control cohort, possibly due to the small sample sizes in these cohorts.

Therefore, the combination of hemocoagulase with a single antifibrinolytic agent appears to be a promising adjuvant strategy for reducing rebleeding risk in aSAH interventions. Nonetheless, this regimen did not positively impact functional outcomes during hospitalization, highlighting the need for future studies to evaluate the long-term outcomes associated with this combination regimen.

### The use of hemostatic drugs alters the incidence of IICP and pulmonary infections following aSAH interventions

ASAH leads to IICP, which subsequently reduces cerebral blood flow, thereby inducing severe complications such as brain metabolic disorders and delayed cerebral ischemia, which contribute to EBI. IICP primarily occurs 3-4 days post-aSAH, underscoring the critical importance of early intervention. Our study revealed that patients treated exclusively with hemocoagulase, rather than those receiving antifibrinolytics alone or in combination with hemocoagulase, exhibited a lower incidence of IICP compared to those did not receive hemostatic agents. This suggests potential neuroprotective effects of hemocoagulase against brain injury following aSAH interventions.

Pulmonary infections (POPs) represent another major complication following aSAH intervention, with an incidence rate ranging from 13% to 29%[34]. POPs are associated with increased postoperative mortality and prolonged hospital stays[35]. Clinical grading scales for assessing SAH severity, such as the World Federation of Neurosurgical Societies (WFNS) grading and the modified Fisher Scale (mFS), also serve as predictive factors for POPs after neurosurgical interventions[34–36]. In this study, we explored the correlation between hemostatic medications and POPs following aSAH interventions. The findings indicated that the use of hemocoagulase alone or in conjunction with a single antifibrinolytic agent was associated with an increased incidence of POPs. Consequently, caution is advised when employing hemocoagulase, given its potential to increase the risk of POPs in aSAH patients.

## Conclusion

Hemostatic agents are used in clinical practice to prevent rebleeding following invasive treatments of aSAH. In our study analyzing data from patients who underwent neurosurgical interventions at three medical institutions over the past decade, we identified that the perioperative use of hemostatic agents is associated with a reduced risk of postoperative neurological deficits. Specifically, combining hemocoagulase with a single antifibrinolytic agent significantly decrease early rebleeding within 72 hours and overall rebleeding, without increasing ischemic risk. Despite this, no significant improvement in overall patient prognosis was observed. The findings suggest that hemostatic medications may improve functional recovery in aSAH patients post-intervention, and the combination of hemocoagulase with a single antifibrinolytic agent effectively lower the risk of early rebleeding. This insight provides valuable guidance for the perioperative application of such medications in the management of aSAH patients undergoing neurosurgical interventions.

Although our study represents the first exploration into the impact of hemocoagulase, both independently and in combination with antifibrinolytics, on overall outcomes and adverse events following aSAH interventions, its retrospective design and limited early case neurological function scores highlight the need for more rigorous future studies. Prospective research is essential to validate these preliminary results and further clarify the optimal use of hemocoagulase-based hemostatic medications for improving patient outcomes.

## Data Availability

All data produced in the present study are available upon reasonable request to the authors.

## Abbreviations

AHA/ASA: American Heart Association/American Stroke Association
aOR: adjusted odds ratios
aSAH: Aneurysmal subarachnoid hemorrhage
CI: Confidence intervals
CTA: computed tomography angiography
CVS: Cerebral vasospasm
DCI: Delayed cerebral ischemia
DSA: digital subtraction angiography
EBI: early brain injury
GCS: Glasgow Coma Scale
H&H: Hunt and Hess
IICP: increased intracranial pressure
LOS: length of stay
MRA: magnetic resonance angiography
mRS: modified Rankin Scale
RIA: ruptured intracranial aneurysm
SD: standard deviation
OR: Odds ratios
POPs: pulmonary infections.

## CRediT authorship contribution statement

**Sijie Sun:** Data curation, Formal analysis, Investigation, Methodology, Writing-original draft. **Jun Ma:** Resources, Methodology. **Jiayi Li:** Resources. **Zhen Liu:** Resources. **Sitong Cui:** Resources. **Qingquan Li:** Investigation. **Hongwei Fan:** Conceptualization, Funding acquisition, Project administration, Validation, Writing-review & editing. **Runze Qiu:** Conceptualization, Project administration, Supervision. **Yingbin Li:** Resources, Project administration, Supervision.

## Conflict of interest

The authors declare that they have no known competing financial interests or personal relationships that could have appeared to influence the work reported in this paper.

## Ethics Declarations

This study was reviewed and approved by the ethics committees of Nanjing First hospital, Nanjing Medical University [KY20230424-01-KS-01], The Second Affiliated Hospital of Nanjing Medical University [2023-KY-147-01], and Sir Run Run Hospital, Nanjing Medical University [2023-SR-020]. Given the retrospective nature of the research and in accordance with institutional guidelines, the requirement for informed consent was waived.

## Acknowledgement

This research was supported by CHANGZHOU SIYAO Hospital Pharmacy Research Fund, Nanjing Pharmaceutical Association (2022YX011) and Nanjing Health Science and Technology Development Special Fund, Nanjing Health Commission (YKK23118).

---

**Supplementary Table 1.**
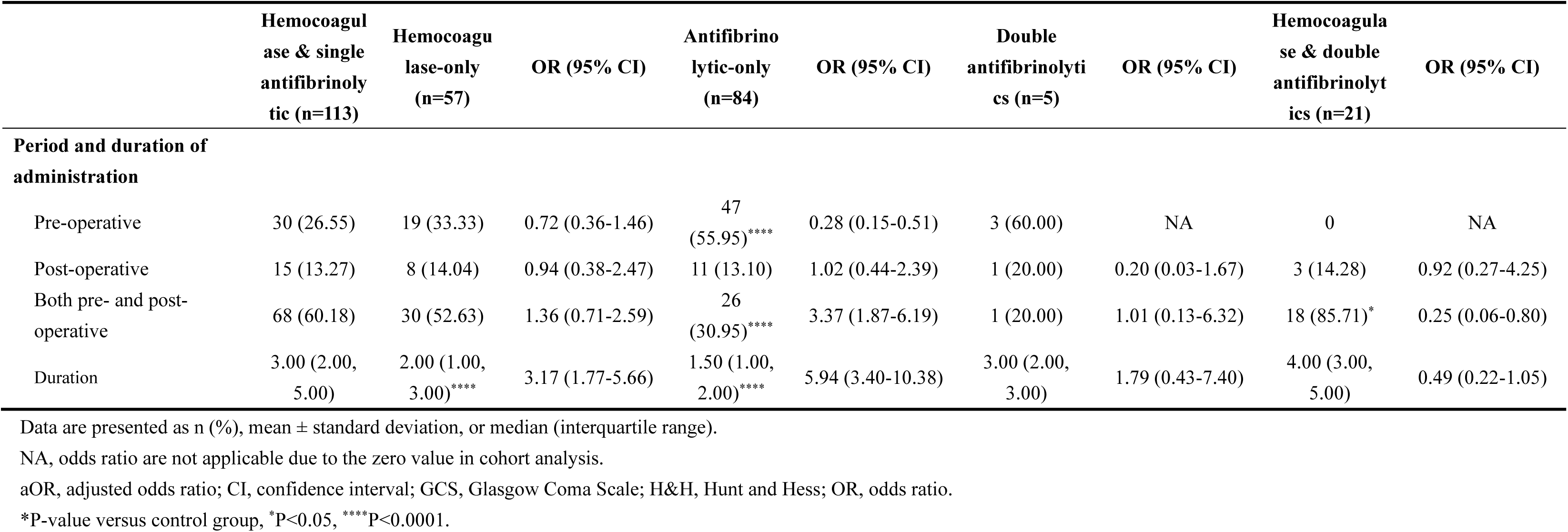
Baseline characteristics comparing cohorts administered hemocoagulase, single antifibrinolytic agents, and other hemostatic strategies.

|  | Hemocoagulase & single antifibrinolytic (n=113) | Hemocoagulase-only (n=57) | OR (95% CI) | Antifibrinolytic-only (n=84) | OR (95% CI) | Double antifibrinolytics (n=5) | OR (95% CI) | Hemocoagulase & double antifibrinolytics (n=21) | OR (95% CI) |
| --- | --- | --- | --- | --- | --- | --- | --- | --- | --- |
| **Period and duration of administration** |  |  |  |  |  |  |  |  |  |
| Pre-operative | 30 (26.55) | 19 (33.33) | 0.72 (0.36-1.46) | 47 (55.95)**** | 0.28 (0.15-0.51) | 3 (60.00) | NA | 0 | NA |
| Post-operative | 15 (13.27) | 8 (14.04) | 0.94 (0.38-2.47) | 11 (13.10) | 1.02 (0.44-2.39) | 1 (20.00) | 0.20 (0.03-1.67) | 3 (14.28) | 0.92 (0.27-4.25) |
| Both pre- and post-operative | 68 (60.18) | 30 (52.63) | 1.36 (0.71-2.59) | 26 (30.95)**** | 3.37 (1.87-6.19) | 1 (20.00) | 1.01 (0.13-6.32) | 18 (85.71)* | 0.25 (0.06-0.80) |
| Duration | 3.00 (2.00, 5.00) | 2.00 (1.00, 3.00)**** | 3.17 (1.77-5.66) | 1.50 (1.00, 2.00)**** | 5.94 (3.40-10.38) | 3.00 (2.00, 3.00) | 1.79 (0.43-7.40) | 4.00 (3.00, 5.00) | 0.49 (0.22-1.05) |

Data are presented as n (%), mean ± standard deviation, or median (interquartile range).
NA, odds ratio are not applicable due to the zero value in cohort analysis.
aOR, adjusted odds ratio; CI, confidence interval; GCS, Glasgow Coma Scale; H&H, Hunt and Hess; OR, odds ratio.
\*P-value versus control group, \*P<0.05, \*\*\*\*P<0.0001.

## Notes

### Competing Interest Statement

The authors have declared no competing interest.

### Funding Statement

This research was funded by CHANGZHOU SIYAO Hospital Pharmacy Research Fund, Nanjing Pharmaceutical Association (2022YX011) and Nanjing Health Science and Technology Development Special Fund, Nanjing Health Commission (YKK23118).

### Author Declarations

Ethics committees of Nanjing First hospital, Nanjing Medical University [KY20230424-01-KS-01], The Second Affiliated Hospital of Nanjing Medical University [2023-KY-147-01], and Sir Run Run Hospital, Nanjing Medical University [2023-SR-020] gave ethical approval for this work.

